# Automated Production of Research Data Marts from a Canonical Fast Healthcare Interoperability Resource (FHIR) Data Repository: Applications to COVID-19 Research

**DOI:** 10.1101/2021.03.11.21253384

**Authors:** Leslie A Lenert, Andrey V. Ilatovskiy, James Agnew, Patricia Rudsill, Jeff Jacobs, Duncan Weatherston, Kenneth Deans

## Abstract

**Objective:** Objective: The COVID-19 pandemic has enhanced the need for timely real-world data (RWD) for research. To meet this need, several large clinical consortia have developed networks for access to RWD from electronic health records (EHR), each with its own common data model (CDM) and custom pipeline for extraction, transformation, and load operations for production and incremental updating. However, the demands of COVID-19 research for timely RWD (e.g., 2-week delay) make this less feasible.

**Methods and Materials:** We describe the use of the Fast Healthcare Interoperability Resource (FHIR) data model as a canonical model for representation of clinical data for automated transformation to the Patient-Centered Outcomes Research Network (PCORnet) and Observational Medical Outcomes Partnership (OMOP) CDMs and the near automated production of linked clinical data repositories (CDRs) for COVID-19 research using the FHIR subscription standard. The approach was applied to healthcare data from a large academic institution and was evaluated using published quality assessment tools.

**Results:** Six years of data (1.07M patients, 10.1M encounters, 137M laboratory results), were loaded into the FHIR CDR producing 3 linked real-time linked repositories: FHIR, PCORnet, and OMOP. PCORnet and OMOP databases were refined in subsequent post processing steps into production releases and met published quality standards. The approach greatly reduced CDM production efforts.

**Conclusions:** FHIR and FHIR CDRs can play an important role in enhancing the availability of RWD from EHR systems. The above approach leverages 21^st^ Century Cures Act mandated standards and could greatly enhance the availability of datasets for research.

## Introduction

The COVID-19 pandemic has illustrated the need for reliable rapidly accessible data from electronic health record (EHR) systems for research on risk factors, predictive models, and evaluation of emerging diseases. Moreover, the lack of reliable large data sets has led to spurious research findings early in the COVID-19 [1]. Two of the largest consortia leverage existing infrastructure for shared data collaboration. The National COVID Collaborative Cohort (N3C) [2] is an alliance among Clinical and Translational Research Grant Awardees sponsored by the National Center to Advancing Translational Science (NCATS). This network leverages past experiences and infrastructure from the Accrual to Clinical Trials Network [3]. N3C’s preferred data model for accepting results is the Observational Medical Outcomes Partnership (OMOP) model maintained by the Observational Health Data Sciences and Informatics (OHDSI) collaborative [4]. However, N3C accepts data in a variety of formats. A second consortium is based around the Patient-Centered Outcomes Research Network (PCORnet) [5] and leverages prior investments on comparative effectiveness research across this large research network [6] to create its database. There also are private large research networks, for example, TriNetX [7], that maintain a large data network of patients with COVID-19 for research from its clinical trial eligibility network [8]. The FDA maintains several large networks for safety evaluation of drugs and devices that are also being applied to problems identified during the pandemic [9]. In addition, some of the same partners in N3C are also using the integrating informatics for integrating biology and bedside (i2b2) platform [10] to study COVID-19. Many networks have overlapping membership and, as a result, members have to maintain duplicative data production processes.

As the pandemic has evolved rapidly, so have the requirements for rapid data updating in these large networks. Minimizing the lag between production of the data through the care of patients using an EHR system and its availability for research increases the relevance of the network to the evolving set of problems seen with COVID-19. In the prior modes of operation, MUSC’s ACT and PCORnet data networks could be used for data operations with lags of three or more months for production, and cycles for new releases of data sets every quarter. In the COVID era, the specifications for the N3C network call for 2-week production cycles for data releases and one-month lags between the closure of a record and its availability within the network. More current data might be even more valuable as new variants of the virus and new therapies emerge.

This is a challenging task that requires the automation of processes for analytic database production.

Production of data for each network is, in itself, a multistep pipeline process that involves mapping and transformation of data to the preferred data model of a research network. Work for data production for different networks is often done in parallel, which is logistically challenging and consumes limited resources. Sometimes work is done in series, mapping from source data to one data model, and then another, which could potentially result in a loss of data through compression or inaccuracies in mapping. In this paper, we describe the use of the Fast healthcare interoperability Resource (FHIR) standard data model as a *canonical model* for initial storage of the data for subsequent transformation to more analytics-oriented models (OMOP and PCORnet) as well as an architecture for multiple simultaneous largely automated translations from FHIR to these two CDMs. This is a particularly important task as the 21st Century Cures Act [11] will require availability from EHRs in FHIR standards for the United States Core Data for Interoperability standards [12], which, while evolving, already has many of the required elements for research CDMs.

## Methods

The approach taken to standardize a data production pipeline for multiple analytic CDMs from FHIR builds on one of the central tools for the FHIR paradigm: a clinical data repository (CDR) designed to store, persist, retrieve, and deliver FHIR resources. A widely used implementation for this operation is the open-source HAPI FHIR engine [13]. We build our system based on the Smile CDR platform [14] that is powered by the HAPI FHIR engine. This platform can accept data in a variety of formats: JSON or XML encoded FHIR objects, HL7 V2.X messages, flat comma-delimited files, and transform these data elements to FHIR resources that are stored in a proprietary relational format, for efficient search and retrieval. Alternatively, some vendors persist FHIR resources using a “data lake” approach, with extensive indexing but a minimal transformation of the data [15].

A standards-specified feature of FHIR CDRs is automated tooling to allow *subscriptions* to specific FHIR resources [14,16]. Subscriptions in the FHIR standard are triggers attached to FHIR data resources. Creating or updating a resource triggers a function that allows copying and transmission of the resource data object to another system. A common use for subscriptions in the FHIR standard is for notification of events. For example, if a patient is registered in an emergency department, a new FHIR resource with the registration information is created, and this then triggers sending a copy of the FHIR resource to another system via FHIR API with JSON payload or other interoperability protocol. This results in the second computer system becoming “aware” of the notification of registration.

We adapted this feature for use in data transformation in our “Federated on FHIR (F-on-F)” architecture [17]. Whenever a new FHIR data resource is created in the clinical FHIR repository, in our system, a copy of the resource is created in a second linked FHIR repository using the subscription mechanism. However, rather than persisting the object in the proprietary database format of the vendor, we co-develop with Smile CDR rule-based transformations implemented in Java to persist data in two different targeted analytical models, OMOP and PCORnet. This results in a system of linked CDMs updated within milliseconds. The approach is illustrated in Figure 1. Both OMOP and PCORnet models are extended to deal with identified data and data elements not covered by the CDM specifications. These are kept in separate tables to preserve the functioning of CDM quality inspection and analytics software.

**Figure 1.**
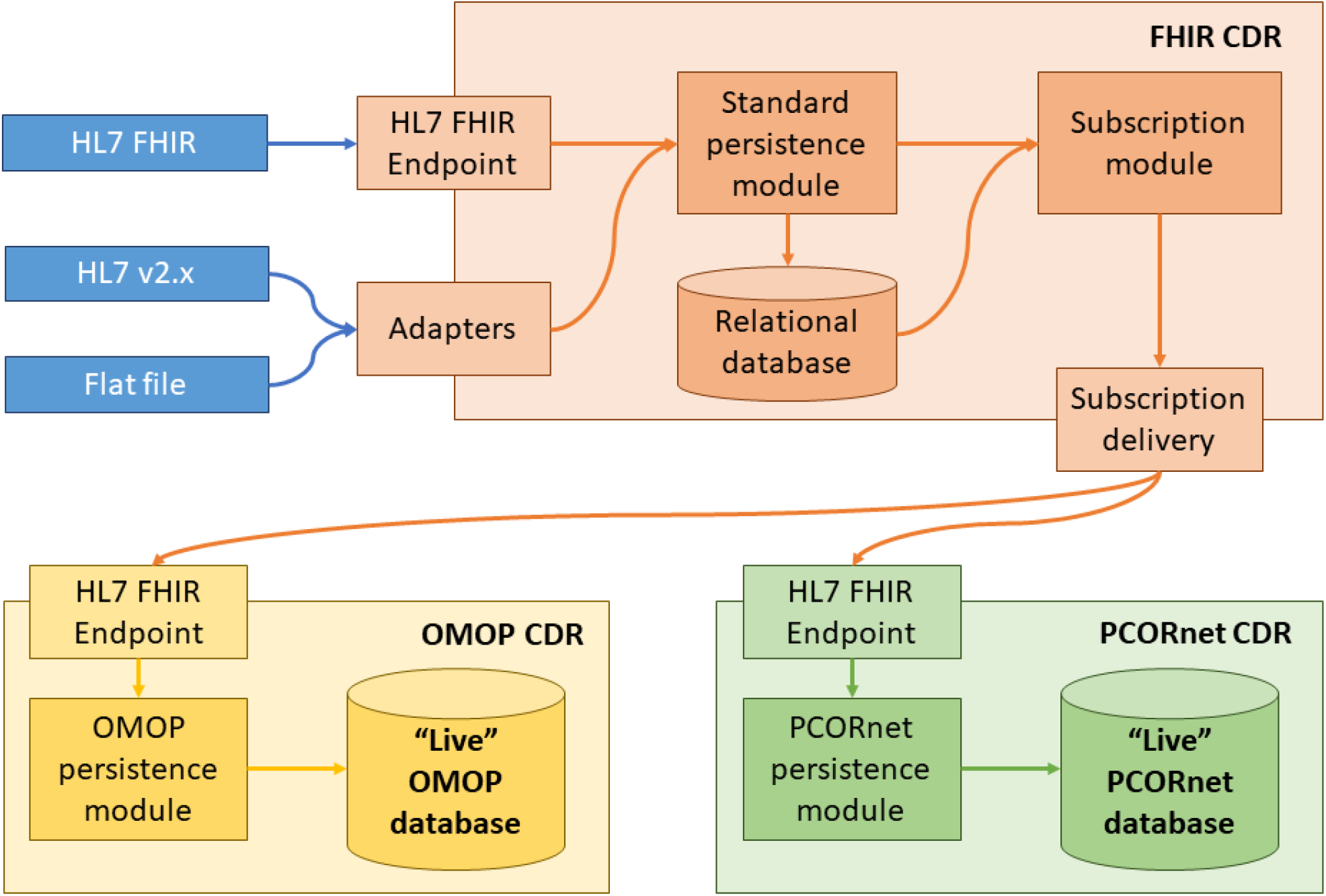
Adapting a FHIR CDR for real-time ETL to OMOP and PCORnet CDMs.

The real-time transformation to analytic CDMs poses additional obstacles due to the transactional nature of the EHR data that evolves and expands for days or even weeks after a given patient encounter, while analytic data models assume a static self-consistent set of data. For example, the OMOP assumes that each patient has a date of birth (known at least with a year precision); in the EHR data the demographics details might be missing and such patients should be removed from the OMOP instance.

Another example, in the OMOP model both visits and associated clinical facts have patient IDs, allowing transitivity violations that occur in the EHR data due to patient merges. If data are incomplete or inconsistent at some point in time, and evolve to completeness and correctness as time passes, the data in the live CDM instances are kept in sync with the source and all updates will be propagated via pipeline. The issue is minimal for study feasibility queries, however, for longitudinal data analysis these inconsistencies need to be resolved. The solution we adopted was a production pipeline with separate static OMOP and PCORnet instances for postprocessing to reconcile the data.

Another issue in design was maintaining the robustness of the process to evolutionary changes in the CDMs. Ongoing changes in CDMs are the rule rather than the exception. The OHDSI group releases a new set of OMOP vocabularies weekly, with changes ranging from adding a few new concepts to complete redesign of the domain organization. Many other networks require frequent vocabulary updates (e.g. once a month for N3C). In case of a major change, the post-processing using an Extract Transform and Load (ETL) approach implemented via SQL was flexible enough to accommodate rapid changes in vocabularies, in contrast to the Java code transformations used in subscription-based mappings. Essentially, the post-processing allows a quasi-incremental approach to the vocabulary updates: the clinical tables built with the old vocabularies are combined with the fresh set of vocabularies and all deviations from the new vocabularies are corrected, with the majority of the data being untouched. This flexibility requires preserving enough “rawness” of the clinical data so updates to an analytic CDM do not require a complete rebuilding of the database. The drawback is that “live” versions of the database produced through subscriptions cannot be used for complex analyses without post-processing (although, again, quick study “feasibility” queries, such as counts of patients with specific e-phenotypes, are possible).

To test the feasibility of this architecture, we converted the Medical University of South Carolina research data warehouse (RDW) and operational production of PCORnet and OMOP CDMs for MUSC to use a novel process based on this concept model. The transformation process and maintenance pipeline was based upon the export of a series of flat pipe-delimited files that were loaded into the FHIR CDR but supports many options for importing EHR data, including HL7 FHIR transactions and HL7 v2.x transactions. In this instance, large export files were then processed to instantiate the repository with pre-existing data. An incremental updating approach based on extracting new data into a flat-file from the RDW was also developed. This incremental update is extracted daily, and loaded into the FHIR repository.

Composite time for database production including the application of post-processing steps in the pipeline was observed. For the OMOP instance, data quality measures were computed using both the in-house reports and OHDSI Achilles and DQD tools. For the PCORnet instance, standardized database quality assessment routines were computed and applied. Results of prior assessments of data quality for PCORnet certification were compared to this new approach for the generation of the database.

## Results

Figure 2. Shows the time delays with different approaches to capture of “raw” EHR data. Flat file exports result in delays of days while data accumulate for export at each stage. Export from our Epic EHR to Epic’s clarity database occurs nightly. Data are extracted from this database daily and stored in a linked clinical CDM based on EHR data model with minimal transformation, and then exported as flat files for conversion to FHIR. HL7 Alternatively, V2.X message feeds from existing data streams can provide near-instantaneous linkages to FHIR CDM assets and result in a near “live” version of PCORnet and OMOP models. These are available for Spartanburg Regional Health System, one of HSSC’s other contributing data partners. Early data products support (blue shading in the figure) support trial feasibility studies (counts); final products support meet network quality requirements and support longitudinal analyses.

**Figure 2.**
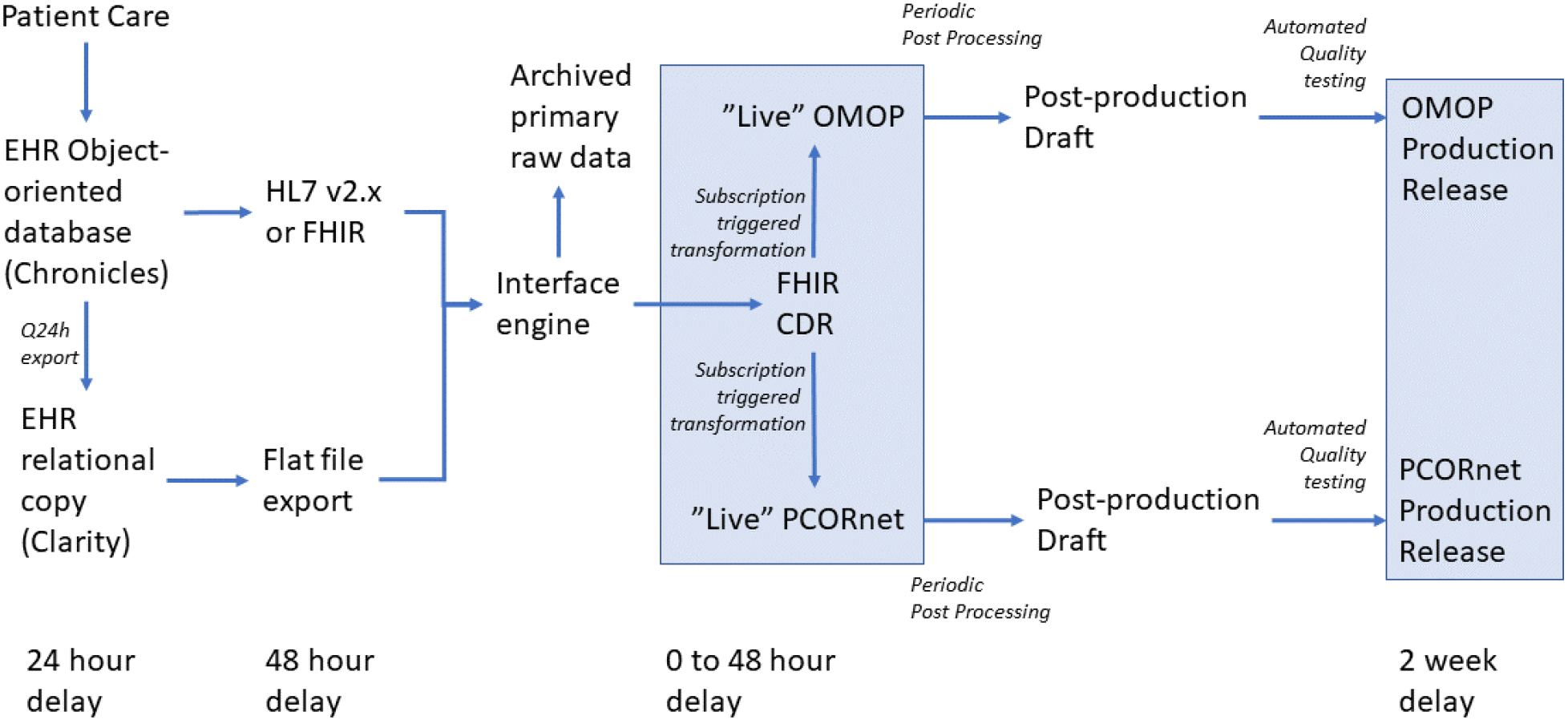
Computational pipeline for simultaneous multi-CDM production. Shaded boxes show available computational products.

Processing for conversion to FHIR, PCORnet and OMOP occurs at HSSC’s Clemson database facility designed to support multiple institutions, each with their own segmented, but linked, FHIR infrastructure.

Table 1 shows the main administrative and clinical data domains implemented in the F-on-F architecture, applied to the MUSC data for the period from 2014-07-01 to 2020-12-31. The results reveal two primary findings. First, the data entry location indicates how the data are represented in each layer. In most cases, there is a 1:1 correspondence between the CDMs, but certain domains are not straightforward. For instance, the vital signs and the lab results are separate domains but exist as common Observation resources in FHIR, stored in both the Measurement and Observation tables in OMOP, and in the individual tables only in PCORnet.

**Table 1.** Comparison of Source RDW, FHIR, OMOP and PCORnet CDMs for MUSC

| Domain | Source | FHIR |  | OMOP |  | PCORnet |  |
| --- | --- | --- | --- | --- | --- | --- | --- |
|  | Count | Resource | Count | Table | Count | Table | Count |
| Patient | 1,078,964 | Patient | 1,063,886 | Person | 1,059,009 | Demographic | 1,063,891 |
| Visit | 10,746,491 | Encounter | 10,636,834 | Visit_Occurrence | 10,628,243 | Encounter | 10,636,928 |
| Diagnosis | 18,402,862 | Condition | 17,593,910 | Condition_Occurrence | 14,254,546 | Diagnosis | 17,594,043 |
| Procedure | 23,029,835 | Procedure | 22,246,999 | Procedure_Occurrence | 16,838,402 | Procedures | 22,247,280 |
| MedOrder | 37,948,002 | MedicationRequest | 32,693,681 | Drug_Exposure | 31,829,609 | Prescribing | 32,703,095 |
| MedAdmin | 84,450,696 | MedicationAdministration | 43,577,863 | Drug_Exposure | 39,562,252 | Med_Admin | 39,574,769 |
| Vital | 16,917,012 | Observation | 16,127,660 | Measurement + Observation | 16,128,048 | Vital | 16,127,920 |
| Lab | 137,984,163 | Observation | 124,870,259 | Measurement + Observation | 154,831,038 | Lab_result_CM | 119,992,129 |

Second, the majority of the metrics are highly consistent between all stages of the pipeline. There are about 11M visits for slightly over 1M patients, associated with 18M diagnoses and 22M procedures. The differences between the CDMs are due to several factors. Most of them are common to all domains: there are certain source data entries that are not processed into the pipeline; the OMOP and PCORnet CDMs require certain data entries to be removed; the asynchronous updates between the systems slightly distort the subset selection of the entries, especially at the end of the date range. The most dramatic differences are OMOP-specific due to the domain assignments (e.g. a diagnosis code might be classified as an observation concept) and one-to-many mappings. The extended metrics report is provided in the Supplementary Table S1.

Initial loading of the backlog of 5-years of EHR data into the FHIR instance required several weeks. However, once loaded, the approach resulted in significant improvements in the time required to produce quarterly updates of the PCORnet instance and improved the concurrency of data in refreshes. As shown in Figure 3, preparation time was far less. Significant work was still required in post-processing but now could be focused on data quality.

**Figure 3.**
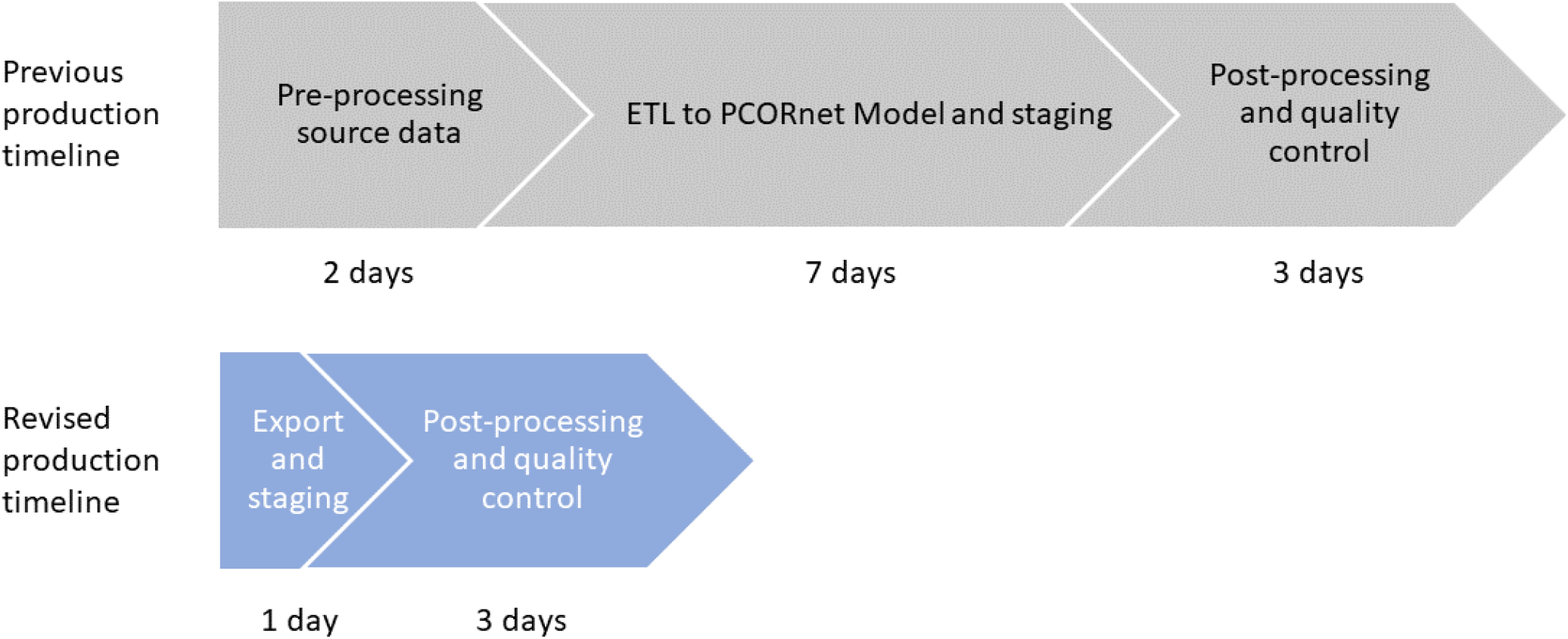
Production timelines for PCORnet database release

## Discussion

The sustainability of research networks for COVID-19 and emerging disorders is an important issue. Costs arise in part due to custom data modeling, ETL tasks, and repetitive data integration tasks required for operations. Further, research infrastructure has to be replicated at each site, requiring significant additional investments. A unique feature of the design is that rather than rely upon the FHIR representation as to the means for query and retrieval of data, the architecture uses FHIR Subscriptions to trigger continuous transformations to other common data models that are maintained in synchrony. This approach also allows the selection of data for specific subsets of patients. Linked databases are available for a query with minimal time lag behind the data source for queries for counts and other simple operations. When an analytical-grade quality of the instance is required, the post-processing can be applied on-demand to produce analytical data sets.

The primary innovations in this work are the use of FHIR as an initial canonical data model and FHIR subscription protocols for the transformation and synchronization of multiple data models. In future work, we will explore the use of subscription models to distribute data across networks and to maintain shared data elements, such as mortality status and social determinants of health data. We will also explore the use of this approach to federate clinical data across sites by maintaining a single master patient identifier and consistent supporting demographic information.

Prior approaches to the problem of maintenance of multiple linked data models in a data repository have focused on other canonical models and *automation* of data both transformation for queries and production of data sets. For example, work from the i2b2 group at Harvard has used the i2b2 data format as the canonical representation to support dynamic ETL from that format to the OMOP and to FHIR [18]. Ong and co-authors use the OMOP model as their canonical representation of data and support dynamic queries mapped from the PCORnet CDM.[19] Choi and colleagues [20] have developed automated mapping functions from OMOP to FHIR for computation.

There is also prior work with the use of FHIR as a meta map for extract transform and load operations between clinical data models. Pfaff and coauthors [21] describe the use of CampFHIR, a tool for guiding ETL for conversions of different models. FHIR concept representation aids and speeds a largely manual ETL process. This approach guides current N3C efforts [2]. The F-on-F approach described herein was developed contemporaneously with CampFHIR [17] with both efforts benefiting from collaborations. F-on-F differs in that it is focused on ongoing data production through automated maintenance of parallel CDM instances. Data transformations are stored/implemented in Java code and implemented in real time. The FHIR CDM servers also provide reference storage and master data management of EHR data and a live canonical representation of the data. The approach may allow us to better understand what is lost in translation.

The use of FHIR as a data source from EHRs is important from a policy perspective as Information Blocking Statutes stemming from the 21^st^ Century Cures Act specify a range of clinical data to be available from EHRs for downloading and data exchange (the USCDI) [11]. They further require that EHRs respond to queries for USCDI elements in the FHIR standard both at an individual patient query level and population level starting in December of 2022. Providers who cannot offer data access in this format may be subject to fines for “information blocking”. EHR vendors must offer these capabilities to maintain certification of their systems as compliant with Meaningful Use regulations. As a result, many barriers for data production for research and safety will be overcome if the starting point for data transformation operations for computational models is the FHIR standard.

The broad future availability of data in the FHIR standard raises the question of whether other analytically oriented models are necessary. OMOP and PCORnet are highly evolved models refined for their purpose [22]. As analytical models, they are optimized for efficient and unbiased analysis of large volumes of longitudinal normalized data. The FHIR model is an object-oriented data model, focused on the accurate expression of clinical events, not computation. F-on-F envisions a best of both worlds approach, with flexible representation and optimized computation.

### Limitations

The above approach is standards-based but leverages proprietary extensions of the FHIR subscription specification. As discussed above, there are inherent limitations in the speed with which clinical data can be integrated into any analytic model such as OMOP or PCORnet. For example, orders or laboratory data cannot be linked to an encounter that does not (yet) exist. The use of the persistence module for the transformation of data is novel and computationally efficient but implements rules in compiled Java code, where changes may be more difficult. Ultimately, some manual ETL was still deemed optimal in the production pipeline; however, future work may reduce these requirements.

### Summary

The use of FHIR standard as a canonical representation of clinical data with the subsequent dynamic transformation to other research CDMs for analytics is a practical approach to accelerate the availability of data for research and may be particularly useful for evolving diseases such as COVID-19. While it is theoretically possible to fully automate transformation to near real-time versions of OMOP or PCORnet databases, it is more practical given the evolving nature of data to take a staged approach for models for longitudinal data analysis applications.

## Data Availability

Data described in this manuscript is available through MUSC's, HSSC's, and PCORnet's data request processes.

## Acknowledgments

This project was supported by grants to Health Sciences South Carolina from the Duke Endowment and by the National Center for Advancing Translational Sciences of the National Institutes of Health under Grant Number UL1 TR001450.

## Notes

### Competing Interest Statement

James Agnew and Duncan Weatherston are employees and own interests in Smile CDR, the FHIR repository described in this study.

### Author Declarations

This study was determined to be a quality improvement study by the Medical University of South Carolina IRB.

